# Generalizing Amyloid *β* Pathology from Postmortem Human Brains with Vision Foundation Models for Lesion Quantification

**DOI:** 10.64898/2026.10.05.26364184

**Authors:** Yuheng Chen, Baayla D. C. Boon, Erika Engelberg-Cook, Lucile Zhu, Darren Rothberg, Annemieke Rozemuller, Dennis Dickson, Melissa E. Murray, Lea T. Grinberg, Daniela Ushizima

## Abstract

**Background:** Traditional neuropathological assessment of Alzheimer’s disease (AD) relies on manual, semiquantitative scoring of amyloid-beta (*Aβ*) pathology. This approach is essential for diagnosis and staging but reduces a continuous and spatially heterogeneous process to ordinal categories and does not capture lesion level variation in plaque and vascular *Aβ*. Amyloid PET enables longitudinal in vivo assessment but it has a limited dynamic range and lacks the sub-millimeter spatial resolution to resolve microscopic plaque morphologies or vascular compartments.

**Methods:** We developed ViTamyloid, a generalizable modular computational pathology pipeline for high throughput segmentation and classification of *Aβ* pathology in digital histological slides of postmortem human brain tissue. In Stage 1, a pathologist supervised framework combining an EfficientNet B4 encoder with a U-Net decoder and a complementary Faster R CNN generated binary segmentation masks and separated confluent lesion candidates. In Stage 2, a frozen high resolution UNI2 h computational pathology foundation model paired with a trainable ViT Adapter classified candidates into five categories: classic cored plaques (CCP), coarse grained plaques (CGP), capillary cerebral amyloid angiopathy (CAA Type 1), parenchymal or leptomeningeal CAA (CAA Type 2), and Diffuse or other *Aβ* positive pathology. We evaluated donor disjoint classification performance, tested generalizability across independent datasets with different antibodies, scanners, resolutions, and image quality, and deployed the pipeline across 1,630 whole slide images from two brain banks.

**Results:** The ViTamyloid achieved a donor disjoint macro F1 of 0.903. AUPRC and AUROC were 0.964 and 0.994 for CCP, 0.947 and 0.990 for CGP, 0.970 and 0.990 for CAA Type 1, and 0.974 and 0.996 for CAA Type 2. The adapted classifier outperformed frozen UNI2 h features alone, which achieved a macro F1 of 0.795. Across independent reference datasets, ViTamyloid showed higher performance than the published pipelines Amyloid-YOLO and PlaqueBox, with nominally significant differences in 26 of 32 paired comparisons.

**Conclusion:** ViTamyloid provides a scalable framework for converting *Aβ* immunohistochemistry into continuous, lesion resolved measurements of plaque and vascular pathology. By preserving lesion identity, morphology, and spatial information, the framework can support studies of AD heterogeneity, clinicopathologic relationships, biomarker concordance, and treatment response.

## 1. Introduction

Alzheimer’s disease (AD) is neuropathologically defined by amyloid-*β* (*Aβ*) plaques and tau pathology. Current diagnostic frameworks integrate the regional distribution of *Aβ*, neurofibrillary degeneration, and neuritic plaques through Thal phases, Braak stages, and CERAD scoring (1–6), while consensus protocols grade cerebral amyloid angiopathy (CAA) on ordinal regional scales (7). These systems are essential for diagnosis and staging, but they intentionally reduce a continuous, spatially heterogeneous process to a small number of ordinal categories. Manual assessment is also time-intensive and varies across observers and laboratories (8–11). Consequently, conventional scores preserve limited information about lesion number, size, morphology, cortical location, and vascular compartment.

This loss of detail matters because *Aβ* deposits are not biologically interchangeable. Classic cored plaques (CCP) are enriched in *Aβ*42 and may occur without cognitive impairment; their clinical significance depends partly on the associated neuritic injury (12, 13). Coarse-grained plaques (CGP) are larger, multicored, *Aβ*40-rich deposits associated with dementia, earlier disease onset, *APOE ε*4 homozygosity, and CAA (12, 14). Although a coarse-grained plaque can superficially resemble a cored plaque, its composition is closer to vascular amyloid, which is why it warrants a class of its own. Vascular *Aβ* represents a separate disease compartment. *Aβ*40 predominates in CAA, contributing to tissue injury through impaired perivascular clearance and vascular dysfunction (15). Capillary involvement (CAA type 1, CAA1) corresponds to a biologically distinct pattern associated with *APOE ε*4, whereas noncapillary vascular amyloid (CAA type 2, CAA2) involves cortical arterioles and leptomeningeal vessels (16, 17). These relationships cannot be reconstructed from total plaque burden or a single ordinal CAA grade.

Lesion-resolved measurement is increasingly important in the anti-*Aβ* treatment era. Clinical trials demonstrate substantial lowering of amyloid PET signal (18, 19), but imaging cannot determine which microscopic deposits were removed, which persisted, or whether *Aβ* shifted among plaques and vessel walls. Autopsy directly resolves lesion identity, vascular compartment, cortical layer, and regional distribution, and can detect sparse residual pathology below the detection threshold of in vivo biomarkers (20–25). Immunotherapy autopsies show that marked parenchymal plaque clearance can coexist with persistent or increased vascular amyloid and pronounced regional heterogeneity (26–34). Quantitative histology is therefore needed to link treatment exposure, amyloid-related imaging abnormalities, biomarkers, and clinical outcomes to the tissue compartments affected.

Brain banks provide the scale needed for this work. Large autopsy cohorts include thousands of donated brains linked to longitudinal clinical, genetic, imaging, and fluid biomarker data (35–37). Yet much of the information in whole-slide images remains inaccessible when pathology is summarized by manual scores. Earlier computational systems showed that convolutional networks and object detectors can recognize broad *Aβ* categories, including cored plaques and CAA (38, 39), and large autopsy studies have demonstrated the value of automated *Aβ* area measurements (40). Their application across brain banks remains constrained by limited lesion taxonomies and by variation in tissue processing, antibodies, staining intensity, scanner optics, resolution, and image quality (41–44). Pathology foundation models may improve portability (45, 46), but they still require task-specific adaptation, dense segmentation, instance separation, artifact rejection, calibration, and neuropathologic expertise.

Here, we developed ViTamyloid (Fig. 1), a modular computational pathology framework that converts *Aβ* immunohistochemistry into continuous measurements of lesion count, area, density, class, and location. The pipeline separates sensitive candidate generation from morphology classification. A segmentation network and a complementary Faster R-CNN (47) resolve *Aβ*-positive components and confluent deposits, while a UNI2-h pathology foundation model (45) with a trainable ViT-Adapter (48) classifies each candidate as CCP, CGP, CAA1, CAA2, or other *Aβ*-positive pathology. We evaluated ViTamyloid using an expert-adjudicated, object-level reference set, deployed it across 1,630 brain bank slides, and tested portability across independent cohorts, antibodies, scanners, and published benchmarks. Our objective was to determine whether the pipeline could retain neuropathologic specificity while generalizing across heterogeneous archival material and recovering continuous biological variation hidden within conventional scores. ViTamyloid is designed not simply to automate plaque detection, but to create a scalable pathomics resource for studying AD heterogeneity, clinical trajectories, biomarker relationships, and treatment response.

**Fig. 1.**
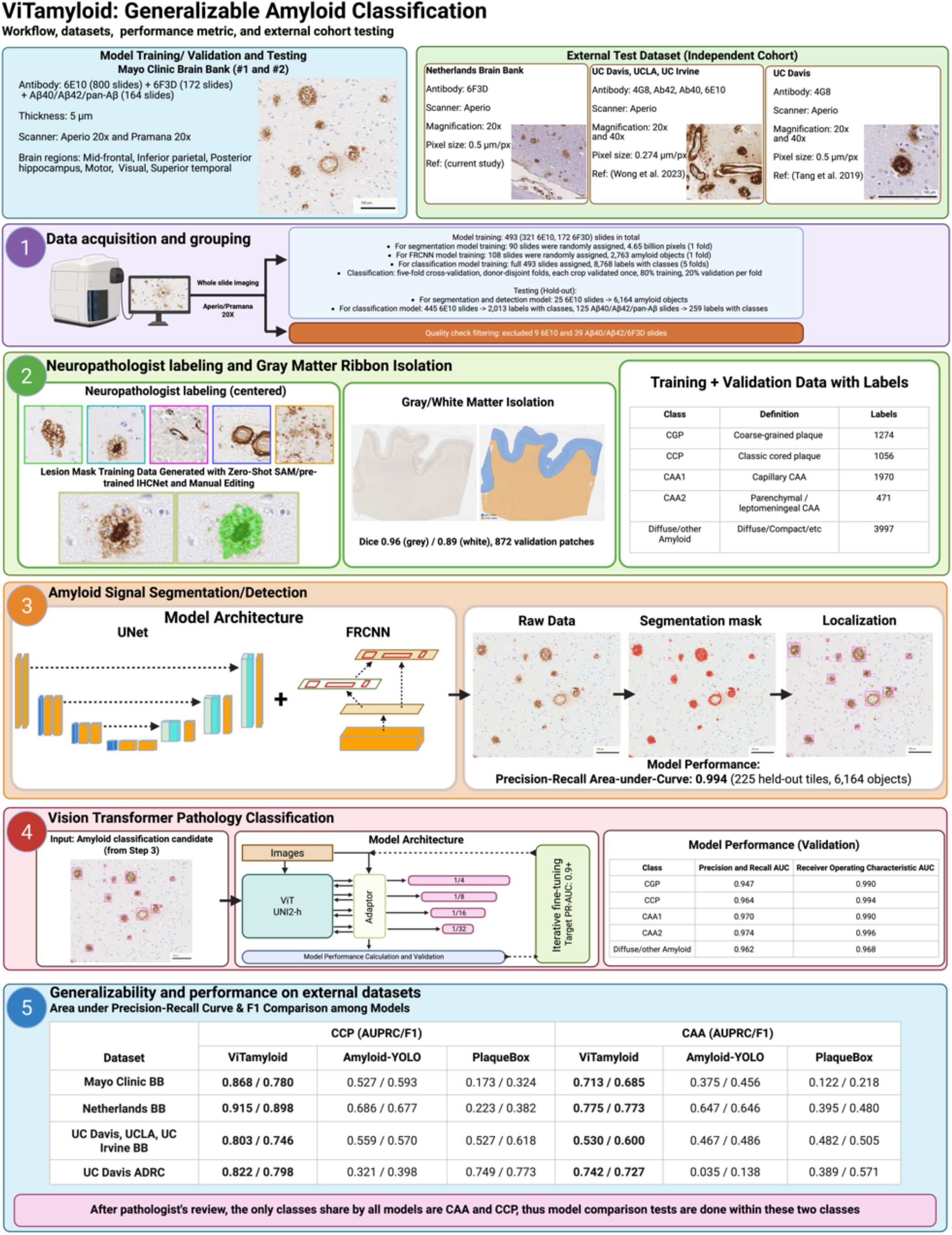
ViTamyloid workflow for converting digitized A*β*-immunostained postmortem brain sections into lesion-level measurements and evaluating performance across internal and external datasets

## 2. Materials and Methods

### 2.1. Study design, cohorts and ethics

Model development and evaluation used archival postmortem brain tissue from the Mayo Clinic Brain Bank (MCBB) and Netherlands Brain Bank (NBB). The MCBB analysis cohorts comprised 1,088 included whole-slide images from 333 unique donors; an additional 48 slides were excluded because of staining or image-quality limitations. The NBB contributed 666 whole-slide images from 97 donors. Cohort composition and clinicopathologic characteristics are summarized in Tables 1 and 2.

**Table 1.**
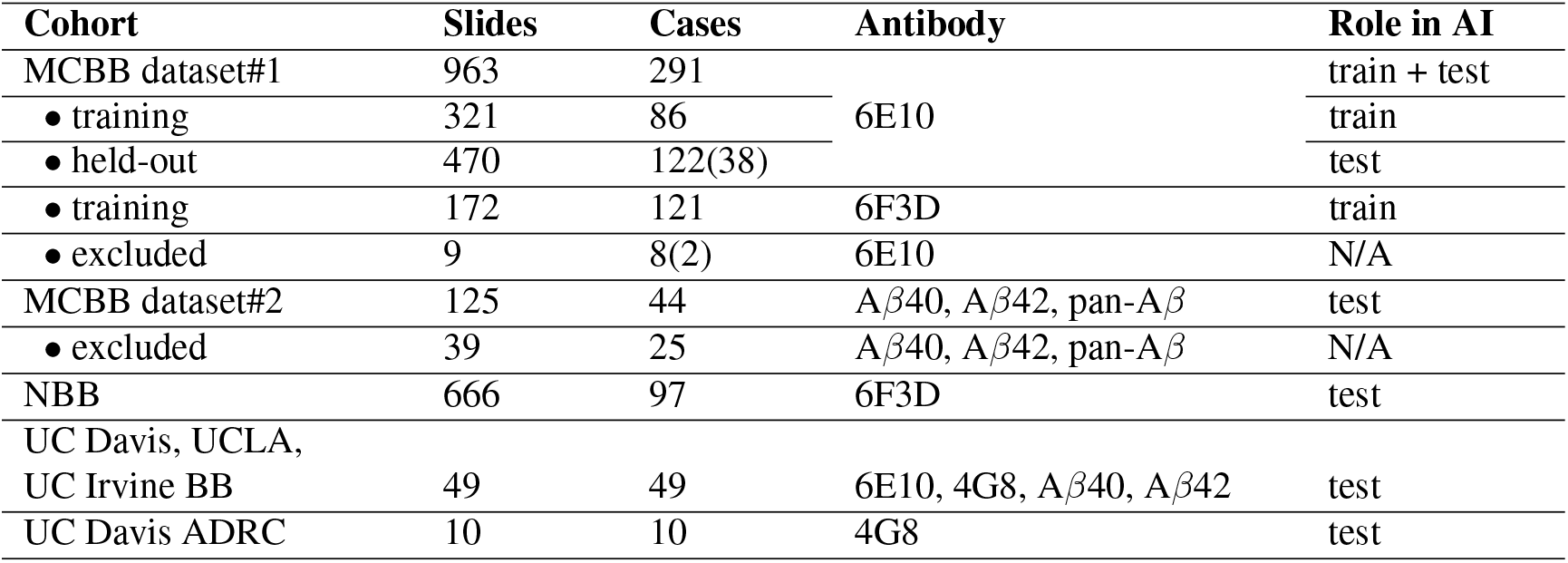
Cohorts in each of the image collections described in Sec. 2. Each collection and subset are slide-disjoint, (#) represent cases that have slides in training dataset as well, since it is possible to have multiple slides from one case. In total, there are 1,630 whole-slide images (MCBB dataset#1 6E10+excluded, MCBB dataset#2+excluded, and NBB), the rest of the external cohorts are cropped images published by the original authors

**Table 2.** Cohort demographics and neuropathology, per case. Continuous variables are median (interquartile range); *n* gives the number of cases with a recorded value where it is not every case. External sets: UC Davis, UCLA and UC Irvine brain banks (Wong et al. (39)) and the UC Davis ADRC hold-out (Tang et al. (38)). N/A: recorded only as free text, not coded. n.r.: not recorded in any available source.

| Variable | Mayo Brain Bank<br>(MCBB #1, 6E10) | Netherlands<br>Brain Bank | Mayo Brain Bank<br>(MCBB #2, test) | Mayo Brain Bank<br>(MCBB #1, 6F3D) | UC Davis, UCLA,<br>UC Irvine BB | UC Davis ADRC<br>hold-out |
| --- | --- | --- | --- | --- | --- | --- |
| Cases | 170 | 97* | 44 | 121 | 49 | 10 |
| Slides imaged | 791 | 666 | 125 | 172 | 49 | 10 |
| Age at death, y, median (IQR) | 67 (62–74) | 75 (68–85) | 74 (66–82) | 82 (75–86) | n.r. | n.r. |
| Sex, M:F | 100:70 | 50:46 | 21:23 | 66:55 | n.r. | n.r. |
| Disease duration, y, median (IQR) | 11 (8–14), *n*=148 | 9 (6–11), *n*=60 | 7 (4–10), *n*=35 | n.r. | n.r. | n.r. |
| APOE ε4 carriers, *n*/*N* (%) | 84/143 (59) | 54/89 (61) | n.r. | 67/106 (63) | n.r. | n.r. |
| Braak stage, median (IQR) | 6 (6–6) | 6 (5–6) | 6 (4–6) | 6 (5–6), *n*=117 | n.r. | n.r. |
| Thal phase, median (IQR) | 5 (5–5), *n*=166 | N/A | 5 (3–5) | 5 (5–5), *n*=102 | n.r. | n.r. |
| CERAD frequent | 137 | N/A | 11 | n.r. | n.r. | 8 |
| CERAD moderate | 8 | N/A | 1 | n.r. | n.r. | 2 |
| CERAD sparse | 6 | N/A | 2 | n.r. | n.r. | 0 |
| CERAD none | 0 | N/A | 1 | n.r. | n.r. | 0 |
| CERAD recorded for | 151/170 | N/A | 15/44 (34%) | 0/121 | – | 10/10 |

\*97 cases contributed slides; 96 have a clinico-pathology record.

- MCBB dataset#1: 791 6E10 slides from 170 donors and 172 6F3D slides from 121 donors. For 6E10: Age, sex, Braak stage and ABC score are recorded for all 170 donors, Thal phase for 166, CERAD for 151 and APOE genotype for 143. This cohort is defined as young-onset AD cohort, because age at onset is under 65 years; it exhibits strongly range-restricted Thal phase and ABC scores (90% of cases with a recorded Thal phase are Thal 5, and 99% of those with a recorded A score are A3). Sampled regions include the mid-frontal, inferior parietal, motor, posterior hippocampus, visual, and superior temporal cortices. For 6F3D: slides come from other MCBB cases outside the young-onset AD cohort: median age at death 82 years (IQR 75–86; 8 died before 65), 55 women and 66 men; Alzheimer’s disease was the primary neuro-pathological diagnosis in 96 (79%), Braak stage median 6 (IQR (inter-quartile range) 5–6; recorded for 117), Thal phase median 5 (recorded for 102), and 67 of the 106 genotyped cases (63%) carry APOE *ε*4.
- MCBB dataset#2: 125 slides from 44 donors (58 pan-A*β* slides from 28 donors, 36 A*β*40 slides from 21 donors and 31 A*β*42 slides from 17 donors, 39 slides are excluded due to poor staining quality). Median age at death is 74 years (IQR 66–82, range 37–95), 23 female and 21 male. Braak stages range from 2 to 6(median 6) and Thal phases from 0 to 5 (median 5), weighted towards advanced disease; CERAD neuritic-plaque scores are recorded for 15 of the 44 donors (34%).
- Netherlands Brain Bank (NBB) (49): 666 6F3D slides from 97 donors (median 7 slides per donor, range 1-8), 96 of whom have a clinico-pathological record. Median age at death is 75 years (IQR 68–85), with 46 women and 50 men; age at onset is recorded for 60 donors (median 68 years, IQR 57–77), giving a median disease duration of 9 years (IQR 6–11). Alzheimer’s disease was the primary neuro-pathological diagnosis in 94 of the 96 donors (98%), which was recorded together with congophilic angiopathy in 5 and with Lewy-body co-pathology in 5, and Braak tangle stage is median 6 (IQR 5–6; recorded for 94). ABC scores, which the NBB holds as free text rather than as graded fields, are available for 83 donors and are as range restricted as those of MCBB dataset #1: A3 in 78, B3 in 72 and C3 in 70. Of the 89 genotyped donors, 54 (61%) carry APOE *ε*4 and 17 are *ε*4 homozygotes; two carry an autosomal-dominant mutation (one APP duplication, one presenilin-1 codon 617) and one a TREM2 variant. Median post-mortem delay is 6.2 h (IQR 4.7–7.0) and median brain weight 1,233 g (IQR 1,080–1,337). Cerebral amyloid angiopathy is documented for 67 of the 96 donors and typed in 56 of those records, with capillary (type 1) in 40 and non-capillary (type 2) in 16, so this cohort’s records resolve the same two CAA classes the pipeline predicts.

To further validate ViTamyloid’s robustness and generalization, and to compare pipelines, we added two external, published slide testing sets for additional model testing and performance comparison to other published pipeline. The prospective validation set of Wong et al. (39) comprises 200 tiles drawn from 49 whole-slide images, one slide per case, across three brain banks and four anti-*Aβ* antibodies: UCLA contributes 26 slides (A*β*40 13, A*β*42 13), the UC Davis brain bank 14 (4G8) and UC Irvine 9 (6E10), and the tiles are balanced at exactly 50 per antibody. Sampling spans the frontal (21 slides), visual (20) and temporal (8) cortices, and the hemisphere is recorded for 26 slides (15 left, 11 right).

Imaging resolution differs by institution, with 0.5 µm per pixel for the UCLA and UC Davis slides against 0.274 µm for those from UC Irvine, and the three sites used three different Aperio instruments (AT2, CS2 and Versa 200), which is the domain shift this set puts to the model. No age, sex, APOE or staging variable is released for these 49 cases.

The PlaqueBox hold-out of Tang et al. (38) is narrower by construction. Every slide is a 5 µm formalin-fixed, paraffin-embedded section of the superior and middle temporal gyrus from the UC Davis Alzheimer’s Disease Center brain bank, stained for 4G8 (residues 17–24, 1:1600, BioLegend SIG-39200) after formic-acid pretreatment and digitized on an Aperio AT2, so the set varies in pathology rather than in region, antibody or hardware. The 10 slides are that study’s hold-out, one slide per decedent case and disjoint from the 33 cases used for their model development. Their recorded CERAD neuritic-plaque score is frequent in 8 and moderate in 2, and the accompanying semi-quantitative temporal-gyrus grades are median 2 for cored plaques (range 1–2) and 3 for diffuse plaques (2–3), while cerebral amyloid angiopathy is graded above zero on only 3 of the 10 slides. Age, sex, APOE and Braak stage are not released, and the published supplement reports NIA-Reagan criteria, Thal phase and CAA type for the development cases rather than for this hold-out.

#### Ethics

This study used deidentified archival postmortem tissue, histologic images, and associated data. The Mayo Clinic determined that this activity did not constitute human subjects research.

### 2.2 Tissue processing, immunohistochemistry and digitization

Formalin-fixed, paraffin-embedded tissue sections were sliced at 5 *µ*m. Tissue sections were processed across institutions using different A*β* antibodies and digitization platforms to evaluate the generalization of the model. Tissue sections were processed across institutions with variations in antibodies and digitization hardware to rigorously stresstest the generalizability of the model. The MCBB dataset #1 cohort utilized anti-*Aβ* clones 6E10 (BioLegend 803001) and 6F3D, while the NBB cohort exclusively utilized clone 6F3D. The MCBB dataset #2 cohort featured diverse targets including *Aβ*40, *Aβ*42, and Pan-*Aβ*. Regarding digitization, whole slides from the MCBB dataset #1 and NBB cohorts were scanned on an Aperio AT2 ScanScope (Leica Biosystems) at × 20 magnification, yielding a native resolution of 0.5025 *µ*m/pixel. Conversely, MCBB dataset #2 slides were digitized using a Pramana scanner at × 20 magnification, producing a native resolution ranging from 0.7181 to 0.7310 *µ*m/pixel. Details for the two published test sets are provided in Wong et al. (39) and Tang et al. (38)

### 2.3 Lesion taxonomy and annotation

Our benchmark dataset considers five amyloid-*β* deposit classes anchored in established neuropathological entities (details and citations in Table 3). CCP represents the primary entity used for CERAD scoring, making them directly comparable to previous automated methodologies. CGPs are morphologically distinct plaques associated with earlier disease onset, which remains unresolved by existing automated methods.

**Table 3.** Class definitions in ViTamyloid.

| Class | Definition | Anchor |
| --- | --- | --- |
| CCP | classic cored plaque, with dense compact core and surrounding corona | CERAD in Mirra et al.(3) |
| CGP | coarse-grained plaque with large, granular/vacuolated deposit lacking a single dense core | Boon et al. 2020 (12) |
| CAA1 | capillary CAA, with Aβ in capillary walls | Thal et al. 2002 (type 1) (16) |
| CAA2 | non-capillary CAA, with parenchymal arteriolar and leptomeningeal vessel walls | Thal et al. 2002 (type 2) (16) |

**Table 4.** Composition of the object-level reference dataset. Counts are the matched analysis set used throughout

| Class | Definition | Crops |
| --- | --- | --- |
| CGP | Coarse-grained plaque | 1274 |
| CCP | Classic cored plaque | 1056 |
| CAA1 | Capillary CAA | 1970 |
| CAA2 | Parenchymal / leptomeningeal CAA | 471 |
| Diffuse/other amyloid | Amyloid outside the four definitions | 3997 |
| **Total (493 slides)** |  | **8768** |

Furthermore, the pipeline distinguishes between CAA Type 1 and Parenchymal/Leptomeningeal CAA (CAA Type 2). This split follows Thal’s capillary-versus-non-capillary typology (16), which is biologically motivated by differing APOE associations. CAA Type 2 deliberately merges parenchymal arteriolar and leptomeningeal vessel involvement, as reliably separating them from a single section is extremely difficult given that leptomeninges are variably present and often detached. A fifth category, designated as “Diffuse/other,” absorbs genuine amyloid signals, including diffuse plaques, that are *Aβ*-positive but are excluded from formal CERAD scoring.

#### Annotation protocol

Slides stained with 6E10 or 6F3D were first processed using gray/white matter isolation to extract the cortical gray matter ribbon (Murray et al. unpublished). From these regions, 1,024 × 1,024-pixel tiles were sampled for annotation. Two annotators (B.D.C. Boon and Y. Chen) labeled A*β*-positive objects as CCP, CGP, CAA1, CAA2, or Diffuse/other Amyloid using custom HTML and Python annotation tools. These labeled images are sent to training-crops generation pipeline: From each bounding box, an effective padding: 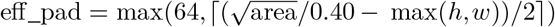 was computed. A square window of side max(*h, w*)+2 · eff_pad, centered on the box and shifted inward at tile edges, was cut and resized to 224 px. The 64 px floor was binding in 94.8 % of the crops for which the parameter was recorded, so **the rule is “bounding box plus 64 px of context per side, resized to 224 px”**: the labels are thus context-normalized rather than lesion-size-normalized.

### 2.4. Dataset construction and partitions

In total, 8,768 mask-paired crops from 493 whole-slide images were annotated: 3,997 Diffuse/other Amyloid, 1,970 CAA1, 1,274 CGP, 1,056 CCP, and 471 CAA2. Of these, 7,893 crops came from 321 MCBB 6E10 slides and 875 from 172 MCBB 6F3D slides. The corpus was expanded through iterative mining and adjudication rather than a prespecified sampling scheme. Diffuse/other Amyloid comprised A*β*-positive deposits that did not meet criteria for CCP, CGP, CAA1, or CAA2, predominantly diffuse plaques. It therefore served as a biologically meaningful residual A*β* category rather than a background class. Primary classifier performance was estimated using five-fold donor-disjoint cross-validation, with all crops and slides from a given donor assigned to the same fold. Every crop was evaluated once in an out-of-fold validation set. No slide contributes crops to both training and validation, every crop is validated once, and validation folds hold 1417–2427 crops (16–28%). Two sensitivity analyses repeat the evaluation with slide-disjoint folds and with the crop-level, antibody-stratified folds, in which crops of one slide can fall in different folds (Supplementary Table 9). No slide used to train in any model contributed to the independent test sets: the 25-slide detection test set and the one-tile-per-slide sets used to compare ViTamyloid with Amyloid-YOLO and PlaqueBox (NBB: 666 slides; MCBB dataset#2: 125 slides), alongside the public sets of UC Davis, UCLA, UC Irvine BB (Wong et al. 2023, 49 slides, 200 tiles) and UC Davis ADRC (Tang et al. 2019, 10 slides, 10,873 crops).

Due to the fact that not all amyloid signals present in each labeled crop are part of the labeled amyloid, a fourth channel is created for each crop and encodes the targeted amyloid binary segmentation with IHCNet (50, 51) with pathologist curated bounding box as object seed, resulting masks are reviewed and edited. These masks enable the isolation of the target amyloid and replace the original background from the crops, becoming an additional augmentation in training stage.

### 2.5. The ViTamyloid pipeline

ViTamyloid processes a whole-slide image in five stages: (i) tissue gating and tiling, (ii) semantic segmentation (50, 51) of amyloid-positive deposits, (iii) candidate detection as the union of segmentation components and a Faster R-CNN (47) proposal set, (iv) five-class classification of each candidate by a two-stream ViT-Adapter (48) ensemble, and (v) classification, and post-processing. Stages (ii)–(iv) are learned; the rest are deterministic.

The design principle throughout is that *proposal and decision are separated*. The segmentation stage is tuned for sensitivity; pathology discrimination is deferred downstream, where two distinct decisions are made by two distinct components. The five-class classifier decides *which morphology* a deposit is, its fifth class being amyloid that meets none of the four named definitions, so all five classes are amyloid and their sum is total amyloid burden. A separate rejection stage decides *whether the object is amyloid at all*, removing blood and vessel-lumen artifact. Folding artifact into Diffuse/other Amyloid would have made that class uncountable; keeping the two decisions apart makes it reportable. Each processing stage in ViTamyloid is further described as follows:

#### (i) Tissue gating and tiling

The down-sampled gray matter mask gates the slide, where the gray/white segmentation model reached Dice 0.96 for gray and 0.89 for white matter on 872 validation patches from 328 slides (patch-level split). Tiles of 1024 × 1024 px are computed on a 768 px grid (25 % overlap) and processed if at least 10 % of the tile is fore-ground; tiles with mean intensity above 242 are discarded as blanks. Because tiles overlap, each candidate is assigned to exactly one tile by a center-in-core rule with a 128 px margin, which removes duplicate detections at tile seams without any later de-duplication.

#### (ii) Resolution normalization

All segmentation, detection and cropping occur in a resampled frame at 0.5025 *µ*m/px, the resolution at which the training dataset was collected. Each tile is read at level 0 as a region of side round(1024*/*scale), where scale = mpp_*x*_*/*0.5025, and resized to exactly 1024 × 1024, for mpp_*x*_ indicating the number of microns per pixel. This allows the pipeline to accommodate different slide-scanner settings: object size in pixels is held comparable to the training configuration.

#### (iii) Detection part 1: Segmentation

Deposits are segmented by a trained MONAI FlexibleUNet (52) model, based on an EfficientNet-B4 encoder (53), trained by distillation from IHCNet: 4,435 tiles of 1024 px sampled from MCBB dataset#1 slides were passed through IHCNet plus its full post-processing chain, and the student was trained on those (image + post-processed mask) pairs with label-smoothed binary cross-entropy, AdamW at learning rate 1e-5, batch 4 on 512 px crops, then specialized for capillary amyloid angiopathy by adding 33 pathologist-curated regions, about 10 percent of the segmentation training dataset. Training stops at epoch 75 with AUCPR at 0.9663. This training was performed in a single full-tile forward pass, taking the foreground softmax probability and thresholding at 0.5. Three deterministic refinements follow. Connected components whose mean hematoxylin optical density exceeds their mean DAB density are rejected after HDX color deconvolution (54). Faint deposit periphery is recovered by a connectivity-constrained DAB expansion, which admits only thresholded regions touching the existing mask; this recovers the periphery without bridging adjacent lesions into one component. Finally, a DAB-guided binary closing (disk radius 6) is applied. Amyloid components are retained at an area between 300 and 200,000 px in size (300 *px*^2^ is approximately 9.8 µm equivalent circular diameter at 0.5025 µm/px).

#### (iv) Detection part 2: Complementary Faster R-CNN (FRCNN) and final amyloid candidates proposal

The final amyloid candidates for the classification stage combine the segmentation components and the FRCNN proposals. **The FRCNN’s role is instance separation, not a second opinion on what amyloid is**. The architecture consists of: ResNet-50-FPN, torchvision; 1,713 pathologist-corrected tiles from 115 slides in the MCBB dataset#1 6E10, sampling specifically confluent amyloid lesion; SGD (lr 2 × 10^*−*3^, momentum 0.9, weight decay 5 × 10^*−*4^), batch 4, up to 30 epochs, slide-level validation split (20%), macro-F1 check-point validation; inference score >= 0.5, NMS IoU 0.3, minimum box 600 *px*^2^. Semantic segmentation labels amyloid pixels but has no notion of objects, so deposits that touch or overlap, such as confluent plaques, or a plaque abutting a vessel wall, emerge as one connected component and would be counted only once by the segmenter alone, possibly causing under-counted quantification. The FRCNN supplies *instance-level* boxes that recover the individual deposits inside such confluent plaque/CAA component.

In detail, each proposal is assigned to a segmentation component by the component contributing the most pixels inside the box, rather than by the label at the box center; this matters for ring-shaped CAA, whose center falls in the lumen (with no amyloid). There are three scenarios in the union-split segmentation postprocessing:

- Where a segmentation component contains **two or more** FRCNN proposal, each proposal is emitted separately and the merged component box suppressed, which *splits* plaques that are confluent;
- Where a segmentation component contains **exactly one** FRCNN proposal, the emitted box is chosen by overlap: at IoU ≥0.5 (same extent) the box will merge, at IoU *<* 0.5 (one a sub-region of the other) the larger is kept;
- Segmentation components claimed by no FRCNN proposal are emitted at their own bounding box.

#### (v) Classification

Each candidate crop is classified by a UNI2-h ViT-Adapter. The frozen branch is the pathology foundation model UNI2-h (45), a ViT-G/14 with depth 24, embedding dimension 1536, SwiGLU MLPs, and eight register tokens, which sees RGB channels and remains frozen throughout (681.4 M frozen parameters). It was trained on a very large archive of histological images, supplies the visual vocabulary, the sense of what stained tissue looks like. The trainable branch (116.7 M parameters) is a ConvNeXt-T (55), in which its task is to extract the fine details of each label and inject into UNI2-h ViT as complementary decision-maker. The decision is then read from the foundation model’s output in two complementary ways: its summary token, and a learned weighted average over all regions of the crop, which lets the classifier weight the parts of the image that matter for the distinction. These are combined and passed through a small two-layer classifier to the five categories: cored, coarse-grained, capillary CAA, parenchymal/leptomenigeal CAA, and Diffuse/other amyloid. Each candidate is assigned with probabilities in each of these 5 classes. Details about the methodology listed below

- **Optimization**: For optimizer, we used AdamW at learning rate 5 × 10^*−*4^, weight decay 0.05, batch size 16, three warm-up epochs then cosine decay, bfloat16 mixed precision, gradient-norm clipping at 1.0, for at most **60 epochs with early stopping after 15 without improvement in validation macro-F1**; Five-fold cross-validation with donor-disjoint folds; random seed 42. Python 3.10, PyTorch 2.8.0 (CUDA 12.8), torchvision 0.23.0, timm 1.0.27, MONAI 1.5.2, scikit-learn 1.7.2, OpenSlide 1.4.3, OpenCV 4.11.0; one NVIDIA L4 24 GB or A100 40 GB per run. The retained checkpoint per fold is the best-macro-F1 epoch. The objective is a focal cross-entropy (*γ* = 2.0) with label smoothing 0.1 and inverse-frequency class weights computed **from the training fold only**. Cut-Mix (*α* = 0.5) is applied to half of batches.
- **Augmentation** follows a specific sequence: geometric transforms (flips, affine *±* 15^*°*^) act on the four-channel tensor; photometric transforms act on RGB only, leaving the mask channel untouched. Two are domain-motivated rather than generic. An **HED stain augmentation** (56) jitters the DAB *colour vector* rather than only its intensity, which allows the model portable across antibodies. A **resolution augmentation** emulating scanners as coarse as 0.85 *µ*m/px and therefore spanning the 0.718–0.731 *µ*m/px of the out-of-domain cohort.
- **Duo-view classifier**: Classifiers of histological objects can learn to read the surrounding tissue rather than the object itself; in pathology image analysis, models trained on unmodified tiles have been shown to over-attend to background and staining noise, and training on inputs whose spatial context is deliberately disrupted forces reliance on intrinsic morphology instead (57). We apply the same principle with an intervention targeted at the labeled object: every training crop is presented twice per epoch, once with its native tissue context and once with all pixels outside the segmentation mask replaced by a gray-matter patch sampled from a reference library. Because the lesion pixels are identical across the two views, a prediction that changes between them can only be driven by the surround, which makes the pair of views a counterfactual test of context-invariance.
- **Inference**. Unlike Liu et al. (57), who infer on unmodified inputs only, ViTamyloid is applied to both original context and disrupted context at inference. For each candidate object, a fixed 224 × 224 px window at 0.5025 µm/px is extracted about the object center and assembled into a four-channel tensor of RGB plus the binary mask of the associated segmentation component. Two forward passes are made, the second after replacing all RGB outside the mask with a sampled gray-matter patch, the draw seeded from the crop’s pixel sum so that an object always receives the same background; the mask channel is identical in both. Softmax outputs are averaged over folds and views, and every result in this paper uses this averaged probability directly: Table 6 assigns each crop its most probable class (argmax), and Table 7 ranks every candidate by its class probability (Section 2.7); no classification threshold is applied.
- **Frozen-feature baseline**. To compare the trained system against its own backbone, frozen UNI2-h class-token features (1,536-d, extracted once over the identical crops) were evaluated with a linear probe (logistic regression, *C* = 1, class-balanced weights) under the identical donor-disjoint folds, preceded by feature standardization. Probe predictions are scored with the same metrics as the pipeline. The probe reads the RGB crop only, so the comparison sets the deployed system as a whole against the foundation model’s frozen representation.

#### Whole-slide deployment and runtime

The pipeline was run end to end on every slide of MCBB dataset#1 (800 slides), the Mayo Clinic Brain Bank collection that contains MCBB dataset#2 (164 slides) and the NBB cohort (666 slides): 1,630 whole-slide images from two brain banks, with no failed run. The MCBB dataset#1 slides were processed with an earlier version of the classifier, the NBB and MCBB dataset#2 slides with the deployed version. Runtime was measured on 15 slides not used in training (five per cohort, chosen at random before timing), each on one NVIDIA A100 40 GB GPU with 8 CPU cores and 80 GB RAM: median 87 min per slide (IQR 58–118 min), 79 min for MCBB dataset#1, 61 min for NBB and 124 min for MCBB dataset#2, scaling with the number of candidates.

### 2.6 Validation: reference standards and performance metrics

#### Pixel-level reference

Segmentation was scored on 110 patches of 1,024 × 1,024 pixels (0.5025 µm per pixel) from 26 slides that supplied no tiles to segmentation training: 37 patches from two NBB slides and 73 from 24 MCBB dataset#1 held-out slides. Patches were centered on amyloid deposits and enriched for difficult boundaries — deposits joined to neighboring material, vessels with a lumen and thin structures — so amyloid occupies 6.7% of their pixels, against a median of 0.31 % of gray matter across whole slides. Every mask was manually edited/created by Y. Chen.

#### Object-level references on our material

To validate Vi-Tamyloid’s robustness and generalization, we evaluated ViTamyloid against highly localized internal ground truths and two independent, publicly available prospective datasets. Object-level detection was evaluated on 25 independent 6E10 slides (225 annotated 1,024 x 1,024 px tiles; 6,164 adjudicated objects). For the comparison with published pipelines, one 1,024 x 1,024 px tile per slide was annotated (by Y. Chen and B. D. C. Boon, reviewed by L. T. Grinberg) on 666 NBB (97 donors) and 125 MCBB dataset#2 (44 donors) slides that contributed to no training stage (tiles with enriched CAA2 and CCP are selected); predictions count only inside these tiles. A prediction only counts toward the evaluation metrics if its centroid falls inside one of these predefined windows.

#### Public benchmarks

The two published sets are described in Section 2.1. On the UC Davis, UCLA and UC Irvine set all 200 tiles are scored against the four-expert consensus boxes (231 cored plaques and 289 CAA; 137 tiles hold at least one consensus object, and detections on the other 63 count as false positives). On the UC Davis ADRC hold-out the 10,873 crops are scored at the authors’ *>* 0.99 vote threshold: 98 crops are cored-positive, in 96 of which the expert marked 97 cores, and 6 crops are CAA-positive.

#### Pixel-level segmentation metrics

The segmentation model is scored per pixel against the 110 manual-edited patches described above, with amyloid pixels as the positive class and all other pixels as negative: the Dice coefficient at the deployed probability threshold of 0.5, and the areas under the precision–recall and receiver operating characteristic curves over all thresholds. This test will answer how well the mask delineates amyloid.

#### Object-level detection metrics

The classification candidate stage, segmentation components with and without the Faster R-CNN proposals, is scored against the 6,164 adjudicated objects of the 25-slide detection test set, a different reference set that asks whether each amyloid deposit is found and separated as one object. Candidates are matched to reference objects by the rule of Section 2.7 (center inside the box or intersection-over-union ≥0.3, one to one in confidence order). We report average precision with candidates ranked by their mean segmentation probability, precision and recall of the permissive candidate pool, and precision and recall at the deployed operating point (minimum segmented area 300 px^2^), the latter also for reference objects of at least 15 µm equivalent circular diameter.

#### Classification metrics

The classifier chooses among five mutually exclusive categories, so a negative class exists for each one and both curves are defined. We report one-class-against-the-rest and pooled over out-of-fold predictions, the area-under-curve of precision-recall curve and the area-under-curve of receiver operating characteristic, together with per-class precision, recall and F1 at the argmax of the averaged probability, macro-F1 across the five classes, accuracy, balanced accuracy (mean per-class recall) and Cohen’s κ; no threshold is applied. Probability calibration is reported as expected calibration error over 15 equal bins of the top-class probability.

### 2.7. External comparators

#### 2.7.1. Comparators and the basis of model/pipeline comparison

Two published brightfield amyloid classification pipelines were run from the authors’ released code and weights (fidelity check: cored AUPRC 0.742 vs 0.743 published) as comparators. Wong et al. (2023) *amyloid-YOLO* is a YOLOv3 detector that emits bounding boxes in two classes, Cored and CAA. Tang et al. (2019) *Plaque-Box* is a three-output multi-label CNN (Cored / Diffuse / CAA) applied to 256-px crops whose locations are proposed by classical HSV brown-blob detection. Both external pipelines/configurations were obtained from the authors’ public GitHub repositories: *keiserlab/amyloid-yolo-paper* and *keiserlab/plaquebox-paper* at, each with the authors’ released weights (*checkpoints/yolov3_ckpt_105*.*pth*; *model-s/CNN_model_parameters*.*pkl*).

Neither comparator has a coarse-grained-plaque class, and neither distinguishes CAA subtypes: each was trained on a single CAA class covering capillary and non-capillary CAA. All comparisons were therefore scored on the two classes that every model was trained on, cored plaque and CAA, using the independent test sets and the scoring rule below.

#### 2.7.2. Scoring rule

Every method was run at its published default configuration and scored by one matching rule, identical for every method on every reference standard: a detection counts as finding a ground-truth (GT) annotated object when either box’s center falls inside the GT or their overlap reaches an intersection-over-union of 0.3, assigned in descending confidence so that each detection claims at most one object and each object is claimed once. The same rule scores the internal detection evaluation (Table 5). Detections/classification are restricted to the annotated region before scoring, and no method/pipeline is scored by the convention of its own paper. Each cell of Table 7 gives the area under the precision– recall curve and the best F1 along that curve, each computed on the method’s own curve, so that no method is penalized for the operating point its authors chose to publish. ViTamyloid enters with each candidate ranked by its class probability (classic cored plaque by its CCP probability; CAA by combining its CAA1 and CAA2 probabilities) to a probability of 0.01; no classification threshold is applied. Detection-s/classification compare only for reference objects of their own class (CAA and CCP/Core plaque, the only two classes shared among the pipelines). CAA is scored as one class on every reference standard, as both comparing pipelines/models were trained on a single CAA class covering capillary and non-capillary CAA: our CAA1 and CAA2 predictions are pooled, reference CAA1 and CAA2 boxes on the NBB and MCBB tiles are both scored as CAA, and CGP boxes are ignore regions whose detections count as neither true nor false positives.

**Table 5.** Object-level detection on the held-out set (25 slides, 225 tiles, 6164 adjudicated objects). **Upper block:** the permissive candidate pool from which the reference was adjudicated, ranked by the mean segmentation probability in each candidate. The Faster R-CNN add 3.4% more candidates and cost 0.004 of precision. **Lower block:** the deployed configuration, which applies a minimum-area gate on top, emits 4215 objects, comprise 4101 from segmentation and 114 from Faster R-CNN. Recall is computed against reference deposits of 15 µm or larger in equivalent circular diameter

| Configuration | Objects | Precision | Recall | AP |
| --- | --- | --- | --- | --- |
| *Permissive pool, no size gate* |  |  |  |  |
| Segmentation alone | 6,015 | 0.991 | 0.967 | 0.966 |
| Union of both sources | 6,219 | 0.987 | **0.996** | **0.994** |
| *Faster R-CNN contribution* | +204 | -0.004 | +0.029 | +0.028 |
| *Deployed configuration* |  |  |  |  |
| Union + minimum-area gate | 4,215 | **0.994** | **0.979** | — |

**Table 6.**
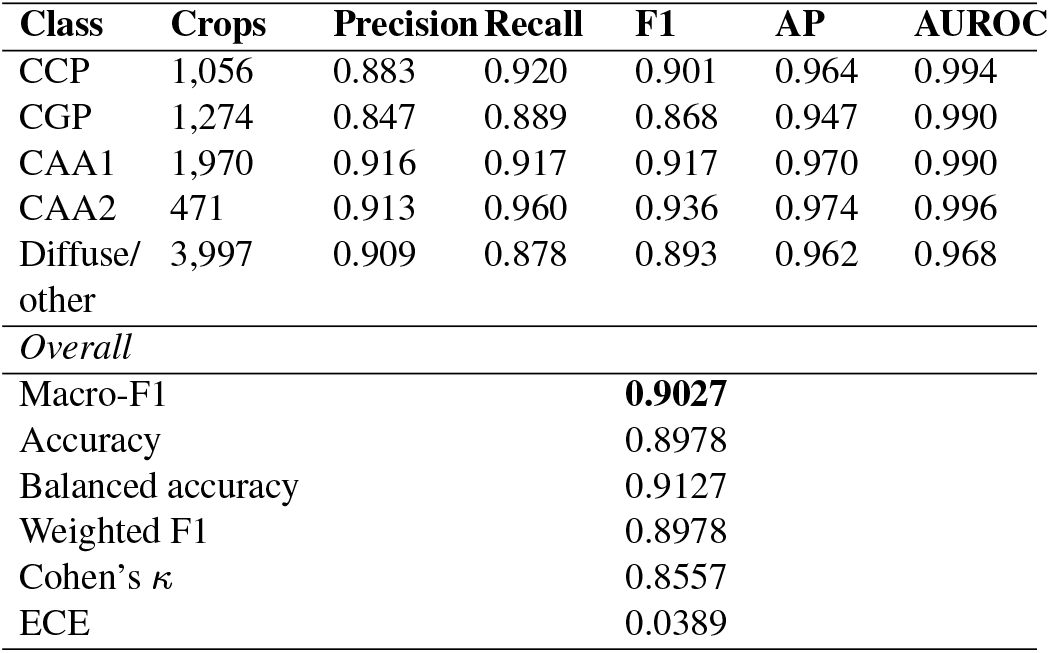
Out-of-fold classification with donor-disjoint folds (the primary analysis: deployed recipe retrained five times, with every young-onset donor’s slides and every slide within one fold), scored through the deployment inference path on 8.768 crops. *AP* is the area under the one-against-the-rest precision–recall curve and *AUROC* the area under the corresponding receiver operating characteristic (Fig. 3); precision, recall and F1 are at the argmax of the averaged probability. 95% confidence intervals (the same recipe with slide-disjoint and crop-level folds) are in Supplementary Table 9.

**Table 7.** Per-class detection performance across four independent reference standards. Each cell gives AUPRC / best F1, computed on each method’s own precision–recall curve so that no method is penalized for the operating point its authors published; precision and recall are evaluated at distinct confidence values. Detections compete only for reference objects of their own class. Every ViTamyloid candidate is ranked by its class probability, every Amyloid-YOLO box by its confidence, and every PlaqueBox object by its predicted class; on the UC Davis ADRC hold-out, PlaqueBox is scored per crop by its class sigmoid, as its authors evaluated it. CAA is scored as one class on every standard: our CAA1 and CAA2 probabilities are summed, reference CAA1 and CAA2 boxes on the MCBB and NBB tiles are both CAA, and CGP boxes are ignore regions charged to no method. On the UC Davis, UCLA and UC Irvine set all 200 expert-reviewed tiles are scored. Best value per standard and class in bold. ^†^ Not significantly different from ViTamyloid (paired bootstrap, *p ≥* 0.05); intervals and tests in Supplementary Tables 10 and 11.

| Reference standard | CCP (AUPRC / best F1) |  |  | CAA (AUPRC / best F1) |  |  |
| --- | --- | --- | --- | --- | --- | --- |
|  | ViTamyloid | Amyloid-YOLO | PlaqueBox | ViTamyloid | Amyloid-YOLO | PlaqueBox |
| Mayo Clinic BB | **0.868 / 0.780** | 0.527 / 0.593 | 0.173 / 0.324 | **0.713 / 0.685** | 0.375 / 0.456 | 0.122 / 0.218 |
| Netherlands BB | **0.915 / 0.898** | 0.686 / 0.677 | 0.223 / 0.382 | **0.775 / 0.773** | 0.647 / 0.646 | 0.395 / 0.480 |
| UC Davis, UCLA, UC Irvine BB | **0.803 / 0.746** | 0.559 / 0.570 | 0.527 / 0.618 | **0.530 / 0.600** | 0.467<sup>†</sup> / 0.486 | 0.482<sup>†</sup> / 0.505 |
| UC Davis ADRC | **0.822 / 0.798** | 0.321 / 0.398 | 0.749<sup>†</sup> / 0.773<sup>†</sup> | **0.742 / 0.727** | 0.035 / 0.138 | 0.389<sup>†</sup> / 0.571<sup>†</sup> |

ADRC: Alzheimer's Disease Research Center

### 2.8. Statistical analysis

The primary estimate of classifier performance is the donor-disjoint five-fold cross-validation (Section 2.4), reported as pooled out-of-fold predictions; the slide-disjoint and crop-level fold designs are sensitivity analyses. Intervals are 95% percentiles of 2,000 bootstrap replicates that resample donors. The slide-disjoint and crop-level fold designs are evaluated on the same replicates, so their differences are paired and judged by the same bootstrap convention as the comparison below (Supplementary Table 9).

For the comparison with published pipelines (Table 7), precision and recall are evaluated at each distinct confidence value, so tied scores enter the curve together, and the area is taken under the interpolated precision envelope. Uncertainty is a 95% percentile interval from 2,000 bootstrap replicates that resample the independent unit of each standard (donor for the MCBB and NBB; slide, one per case, for the UC Davis, UCLA and UC Irvine material and for the UC Davis ADRC hold-out) and re-score the fixed matched detections with those weights. Every method is scored on the same replicates, so the difference between ViTamyloid and each comparator is paired; its two-sided *p*-value is twice the share of replicates on the far side of zero, with *p <* 0.05 taken as significant, and is not adjusted for multiple comparisons (Supplementary Tables 10 and 11).

## 3. Results

### 3.1. An adjudicated, object-level reference dataset

We assembled an expert-labeled reference corpus of 8768 *β* positive lesion crops from 493 whole-slide images for training and cross-validation of the ViTamyloid classifier (Table 4).

**Table 8.** Comparative analysis of plaque detection pipelines.

| Comparative Dimension | PlaqueBox (Tang et al., 2019) | Amyloid-YOLO (Wong et al., 2023) | ViTamyloid (this work) |
| --- | --- | --- | --- |
| **Core Paradigm** | From scratch CNN tile level classification. | Object detection bounding box pipeline. | Decoupled foundation model "pathomics" toolkit. |
| **Backbone Architecture** | Optimized six layer convolutional architecture with two dense layers. | YOLOv3 (Darknet 53 backbone) coupled with a consensus filtering CNN. | Segmentation by a U-Net with an EfficientNet-B4 encoder; classification by a two-stream ViT-Adapter pairing the frozen UNI2-h pathology foundation model (681 M parameters) with a trainable four-channel ConvNeXt-T (117 M). |
| **Detection & Segmentation Approach** | Hue and size based thresholding (LCH/HSV) isolates candidate objects centered on 256x256px tiles. | End to end object coordinates generated by an iterative merging watershed process. | Per-pixel segmentation of DAB-positive deposits yields candidate components; a Faster R-CNN proposal set to separate confluent deposits into instances. |
| **Labeling & Training Effort** | >70,000 manual expert tile annotations across multiple curation phases. | Repurposed sparse datasets with no human drawn bounding boxes. | Zero shot localization/segmentation (SAM) or pretrained IHCNet with minimal need of manual editing. |
| **Plaque Morphologies** | Cored plaques, diffuse plaques, and CAA. | Cored plaques and CAA (diffuse plaques excluded as they lack defined boundaries). | Classic cored (CCP) and coarse-grained (CGP) plaques, and CAA resolved into capillary (Type 1) and parenchymal/leptomeningeal (Type 2); diffuse plaque is retained in a separate Other Amyloid class rather than discarded. |
| **Key Performance Highlights** | Held out set (Phase III) AUROC of 0.993 and AUPRC of 0.743 for cored plaques. | Matched human experts in cored plaque AP (0.64) and surpassed them in CAA (0.75 AP vs. 0.51 AP). | Donor-disjoint out-of-fold macro F1 0.903 over five classes; candidate detection AP 0.994 (recall 0.996); deployed precision 0.994. The only pipeline of the three that resolves coarse-grained plaque and the two CAA subtypes. |
| **Speed & Clinical Scalability** | Sliding window approach requires 3 hours 4 minutes per WSI on a GPU. | Scores a whole slide in 1 min 40s (GPU) or 5 min 30s (CPU) on standard workstations. | Run and scaled across 1,630 whole-slide images from two brain banks. Median 87 minutes per slide (IQR 58–110) on one NVIDIA A100 GPU |

**Table 9.** Detailed performance results summarized in Table 6 (donor-disjoint folds: all slides of a donor fall within one fold) with 95% confidence intervals and the two five-fold sensitivity analyses: 1. Slide-disjoint folds that keep every slide within one fold. 2. Crop-level folds that stratified by antibody at the crop level, so crops from one slide can fall in both training and validation folds. Intervals are percentiles of 2,000 bootstrap replicates that resample donors, with all slides of a donor (MCBB dataset#1 6E10 or 6F3D) kept together; all three designs are evaluated on the same replicates, so their differences are paired. Class assigned by the argmax of the averaged probability; 8,768 crops.

| Folds | Class | Precision | Recall | F1 | AP | AUROC |
| --- | --- | --- | --- | --- | --- | --- |
| Donor-disjoint (primary analysis, Table 6) | CGP | 0.847 (0.75–0.91) | 0.889 (0.82–0.93) | 0.868 (0.79–0.92) | 0.947 (0.88–0.98) | 0.990 (0.98–0.99) |
|  | CCP | 0.883 (0.85–0.92) | 0.920 (0.90–0.94) | 0.901 (0.88–0.93) | 0.964 (0.95–0.98) | 0.994 (0.99–1.00) |
|  | CAA1 | 0.916 (0.89–0.94) | 0.917 (0.88–0.94) | 0.917 (0.89–0.93) | 0.970 (0.96–0.98) | 0.990 (0.99–0.99) |
|  | CAA2 | 0.913 (0.86–0.96) | 0.960 (0.94–0.98) | 0.936 (0.90–0.96) | 0.974 (0.96–0.99) | 0.996 (0.99–1.00) |
|  | Diffuse/other | 0.909 (0.88–0.93) | 0.878 (0.84–0.91) | 0.893 (0.86–0.92) | 0.962 (0.94–0.98) | 0.968 (0.96–0.98) |
| Slide-disjoint (sensitivity analysis #1) | CGP | 0.832 (0.74–0.90) | 0.889 (0.82–0.93) | 0.860 (0.78–0.91) | 0.940 (0.87–0.97) | 0.989 (0.98–0.99) |
|  | CCP | 0.862 (0.83–0.89) | 0.908 (0.86–0.95) | 0.885 (0.86–0.91) | 0.951 (0.93–0.97) | 0.992 (0.99–1.00) |
|  | CAA1 | 0.926 (0.90–0.95) | 0.909 (0.87–0.93) | 0.917 (0.89–0.93) | 0.972 (0.96–0.98) | 0.991 (0.99–0.99) |
|  | CAA2 | 0.915 (0.87–0.95) | 0.960 (0.94–0.98) | 0.937 (0.90–0.96) | 0.975 (0.96–0.99) | 0.996 (0.99–1.00) |
|  | Diffuse/other | 0.906 (0.88–0.93) | 0.876 (0.84–0.90) | 0.891 (0.86–0.91) | 0.961 (0.94–0.97) | 0.966 (0.96–0.97) |
| Crop-level (sensitivity analysis #2) | CGP | 0.882 (0.81–0.93) | 0.860 (0.78–0.91) | 0.871 (0.80–0.92) | 0.945 (0.88–0.98) | 0.989 (0.98–0.99) |
|  | CCP | 0.867 (0.84–0.91) | 0.930 (0.91–0.95) | 0.898 (0.88–0.92) | 0.958 (0.94–0.98) | 0.994 (0.99–1.00) |
|  | CAA1 | 0.930 (0.91–0.94) | 0.930 (0.90–0.95) | 0.930 (0.91–0.94) | 0.977 (0.96–0.98) | 0.992 (0.99–0.99) |
|  | CAA2 | 0.933 (0.90–0.96) | 0.970 (0.95–0.99) | 0.951 (0.93–0.97) | 0.978 (0.96–0.99) | 0.997 (0.99–1.00) |
|  | Diffuse/other | 0.910 (0.89–0.93) | 0.895 (0.87–0.92) | 0.902 (0.88–0.92) | 0.964 (0.95–0.97) | 0.970 (0.96–0.98) |

| Folds | Macro-F1 | Accuracy | Balanced accuracy | $\kappa$ | ECE |
| --- | --- | --- | --- | --- | --- |
| Donor-disjoint (Table 6) | 0.903 (0.88–0.92) | 0.898 (0.88–0.91) | 0.913 (0.89–0.93) | 0.856 (0.83–0.88) | 0.039 (0.03–0.05) |
| Slide-disjoint | 0.898 (0.88–0.91) | 0.894 (0.88–0.91) | 0.908 (0.89–0.92) | 0.850 (0.82–0.87) | 0.050 (0.04–0.06) |
| Crop-level | 0.910 (0.89–0.92) | 0.906 (0.89–0.92) | 0.917 (0.90–0.93) | 0.867 (0.84–0.89) | 0.043 (0.04–0.05) |

Macro-F1 differences: donor-disjoint vs. slide-disjoint +0.0049 (95% CI –0.0016 to +0.0120; paired $p = 0.138$ ); crop-level vs. donor-disjoint +0.0077 (95% CI –0.0029 to +0.0172; paired $p = 0.150$ ); crop-level vs. slide-disjoint +0.0126 (95% CI +0.0032 to +0.0218; paired $p = 0.010$ ).

**Table 10.**
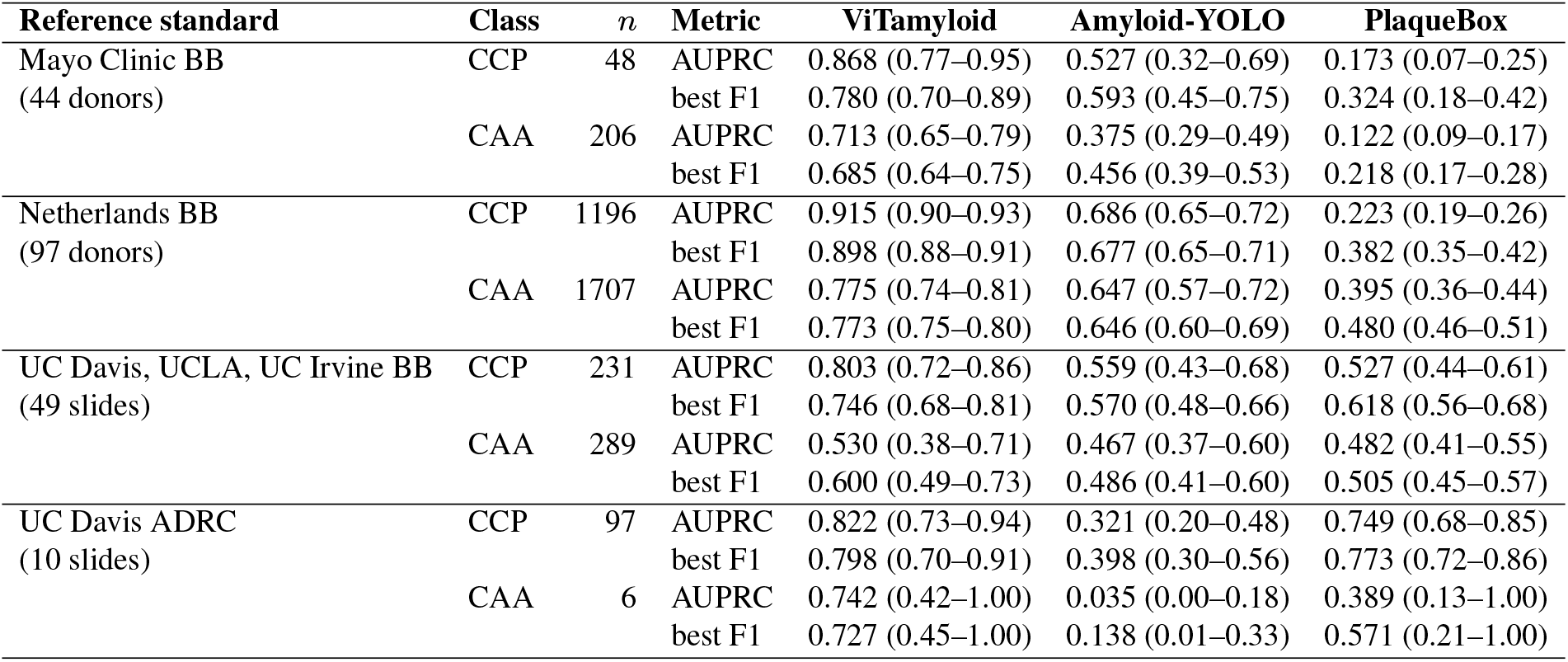
Detailed performance analysis, summarized in Table 7 with 95% confidence intervals. Intervals are percentiles of 2,000 bootstrap replicates that resample the independent unit named under each standard (donor where donor identity is known; slide, one per case, otherwise) and re-score the fixed matched detections; every method is scored on the same replicates. Precision and recall are evaluated at distinct confidence values. *n* = reference objects of the class (UC Davis ADRC CAA: crops).

**Table 11.** Difference between ViTamyloid and each comparator on every cell of Table 7, from the same 2,000 paired bootstrap replicates as Supplementary Table 10. Two-sided *p* is twice the share of replicates on the far side of zero and is not adjusted for multiple comparisons; with 2,000 replicates the smallest attainable value is 0.0005, shown as *<*0.001.

| Reference standard | Class | Metric | Comparator | Difference (95% CI) | $p$ |
| --- | --- | --- | --- | --- | --- |
| Mayo Clinic BB | CCP | AUPRC | Amyloid-YOLO | +0.341 (+0.168 to +0.562) | $<0.001$ |
|  | CCP | best F1 | Amyloid-YOLO | +0.187 (+0.007 to +0.372) | 0.041 |
| | CCP | AUPRC | PlaqueBox | +0.696 (+0.565 to +0.835) | $<0.001$ |
| | CCP | best F1 | PlaqueBox | +0.456 (+0.317 to +0.662) | $<0.001$ |
| | CAA | AUPRC | Amyloid-YOLO | +0.338 (+0.225 to +0.440) | $<0.001$ |
| | CAA | best F1 | Amyloid-YOLO | +0.229 (+0.162 to +0.307) | $<0.001$ |
| | CAA | AUPRC | PlaqueBox | +0.591 (+0.508 to +0.678) | $<0.001$ |
| | CAA | best F1 | PlaqueBox | +0.467 (+0.387 to +0.554) | $<0.001$ |
| Netherlands BB | CCP | AUPRC | Amyloid-YOLO | +0.229 (+0.196 to +0.263) | $<0.001$ |
| | CCP | best F1 | Amyloid-YOLO | +0.220 (+0.191 to +0.248) | $<0.001$ |
| | CCP | AUPRC | PlaqueBox | +0.692 (+0.652 to +0.725) | $<0.001$ |
| | CCP | best F1 | PlaqueBox | +0.515 (+0.475 to +0.556) | $<0.001$ |
| | CAA | AUPRC | Amyloid-YOLO | +0.128 (+0.081 to +0.176) | $<0.001$ |
| | CAA | best F1 | Amyloid-YOLO | +0.127 (+0.095 to +0.159) | $<0.001$ |
| | CAA | AUPRC | PlaqueBox | +0.380 (+0.349 to +0.410) | $<0.001$ |
| | CAA | best F1 | PlaqueBox | +0.293 (+0.266 to +0.316) | $<0.001$ |
| UC Davis, UCLA, UC Irvine BB | CCP | AUPRC | Amyloid-YOLO | +0.244 (+0.145 to +0.339) | $<0.001$ |
| | CCP | best F1 | Amyloid-YOLO | +0.175 (+0.103 to +0.249) | $<0.001$ |
| | CCP | AUPRC | PlaqueBox | +0.275 (+0.206 to +0.334) | $<0.001$ |
| | CCP | best F1 | PlaqueBox | +0.128 (+0.067 to +0.189) | $<0.001$ |
|  | CAA | AUPRC | Amyloid-YOLO | +0.063 (-0.029 to +0.192) | 0.190 |
|  | CAA | best F1 | Amyloid-YOLO | +0.114 (+0.038 to +0.206) | 0.001 |
|  | CAA | AUPRC | PlaqueBox | +0.048 (-0.072 to +0.207) | 0.408 |
|  | CAA | best F1 | PlaqueBox | +0.095 (+0.005 to +0.205) | 0.037 |
| UC Davis ADRC | CCP | AUPRC | Amyloid-YOLO | +0.501 (+0.376 to +0.631) | $<0.001$ |
| | CCP | best F1 | Amyloid-YOLO | +0.400 (+0.286 to +0.519) | $<0.001$ |
|  | CCP | AUPRC | PlaqueBox | +0.073 (-0.063 to +0.212) | 0.352 |
|  | CCP | best F1 | PlaqueBox | +0.024 (-0.109 to +0.158) | 0.703 |
| | CAA | AUPRC | Amyloid-YOLO | +0.706 (+0.359 to +0.993) | $<0.001$ |
| | CAA | best F1 | Amyloid-YOLO | +0.589 (+0.270 to +0.949) | $<0.001$ |
|  | CAA | AUPRC | PlaqueBox | +0.353 (-0.500 to +0.537) | 0.267 |
|  | CAA | best F1 | PlaqueBox | +0.156 (-0.333 to +0.471) | 0.476 |

Labels were accumulated through iterative extraction and adjudication: capillary CAA (CAA1, 1970) and coarse-grained plaques (CGP, 1274) are the most numerous named classes, whereas parenchymal and leptomeningeal CAA (CAA2, 471) is the least. The Diffuse/other Amyloid class comprised *β*-positive deposits that did not meet criteria for CCP, CGP, CAA1, or CAA2. It was therefore a measurable A*β* category rather than a background or artifact class. Total amyloid is therefore the sum of CGP, CCP, CAA1, CAA2 and Diffuse/Other Amyloid. Blood cells, vessel lumen and similar chromogen artifact are removed during detection stage (Section 2.5) rather than being folded into Diffuse/Other Amyloid, so that it remains a countable amyloid category rather than a residual bin for anything unclassified.

### 3.2. Cross-validated ViTamyloid performance

#### 3.2.1. Robust amyloid candidates detection

The detection stage proposes candidates from segmentation components and Faster R-CNN complementary detection. Model performance metrics are scored without class, only “amyloid vs background/noise/debris”, since it emits amyloid classification candidates rather than subtypes.

##### At the pixel level, the segmentation recovers the majority of amyloid lesions

Scored against the 110 hand-annotated patches of Sec.2.6 (73 MCBB dataset#1 held-out and 37 NBB patches from 26 slides; 115.3 million pixels), the Dice coefficient at the default threshold of 0.5 is 0.904 and the area under the precision–recall curve is 0.938 (Fig. 2).

**Fig. 2.**
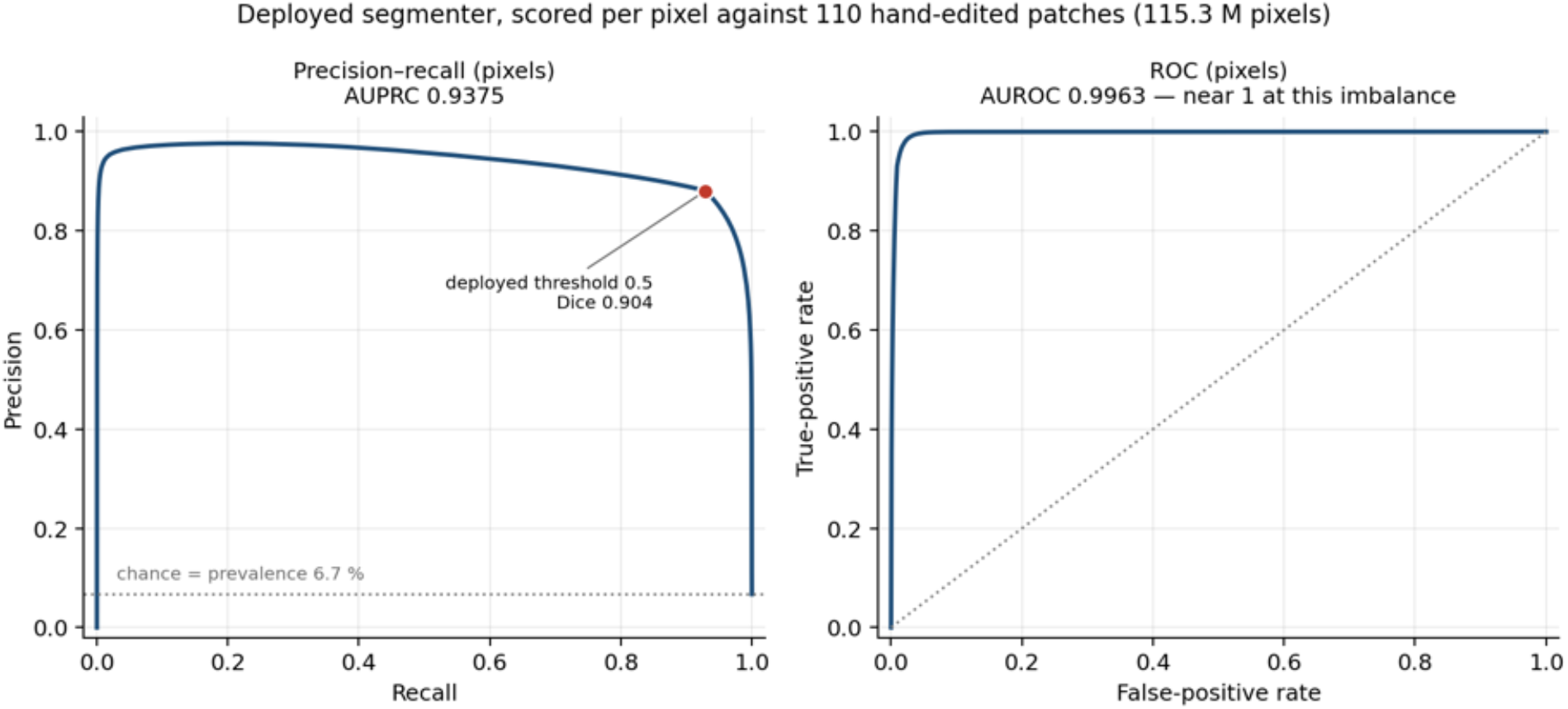
Segmentation scored per pixel against 110 hand-edited patches (26 slides, 115.3 million pixels, 6.7% amyloid). **Left**, precision–recall; the marker is the deployed threshold of 0.5, and the dotted line is chance, which equals prevalence. **Right**, receiver operating characteristic.

##### FRCNN is complementary to the segmentation model

Segmentation alone reaches average precision 0.966 and recall 0.967 (Table 5). Adding Faster R-CNN proposers contributes 3.4% more candidates (204 unique Faster R-CNN candidates) at a low cost of precision (0.004), and raises recall by 0.029 and average precision by 0.028. These additional amyloid candidates are deposits that merge and cannot split by segmentation alone. The union of segmentation and FRCNN is therefore not redundancy, but deposits separation. At the deployed operating point, the pipeline emits 4,215 objects at precision 0.994, recovering 0.979 of reference deposits of 15 µm or more in equivalent circular diameter (about 700 *px*^2^); the deployed minimum-area gate is 300 *px*^2^ of segmented area (about 9.8 µm).

#### 3.2.2. Cross-validated amyloid classification performance

The classifier is a ViT-Adapter: a frozen UNI2-h vision foundation model into which a four-channel ConvNeXt-T adapter injects tokens, read out by an attention-pooled head. Two arms are trained, one on the RGB crop and one on the same crop with its background replaced by sampled gray matter, and their probabilities are averaged. Performance is reported as pooled out-of-fold predictions over 8.768 crops from five-fold cross-validation with donor-disjoint folds, scored through the deployment inference path.

##### The five amyloid lesion classes separate cleanly

With donor-disjoint folds, macro-F1 is 0.903 (95% CI 0.883 to 0.918; accuracy 0.898, balanced accuracy 0.913, Cohen’s *κ* 0.856). As sensitivity analyses, the same recipe gives 0.898 with slide-disjoint folds and 0.910 with the crop-level folds, in which crops of one slide can fall on both sides; neither differs significantly from the donor-disjoint estimate (Supplementary Table 9). Per class, F1 ranges from 0.868 for coarse-grained plaques to 0.936 for parenchymal CAA (Table 6), and one class against the rest gives average precision 0.947–0.974 and area under the receiver operating characteristic 0.968–0.996 (Fig. 3). The two CAA subtypes, which no external method distinguishes, reach F1 0.917 (capillary) and 0.936 (parenchymal), and cored plaques 0.901. The weakest class is coarse-grained at 0.868, and its errors are almost entirely with Diffuse/other amyloid rather than with another amyloid subtype (Fig. 4). Ranked by average precision the order is the same: parenchymal CAA (0.974) > capillary CAA (0.970) > classic cored plaque (0.964) > coarse-grained plaque (0.947). On the same folds, frozen UNI2-h class-token features reach macro-F1 0.795 with a linear probe; the largest gain from the trained classifier is for classic cored plaques (F1 0.901 against 0.754 for the frozen UNI2-h features).

**Fig. 3.**
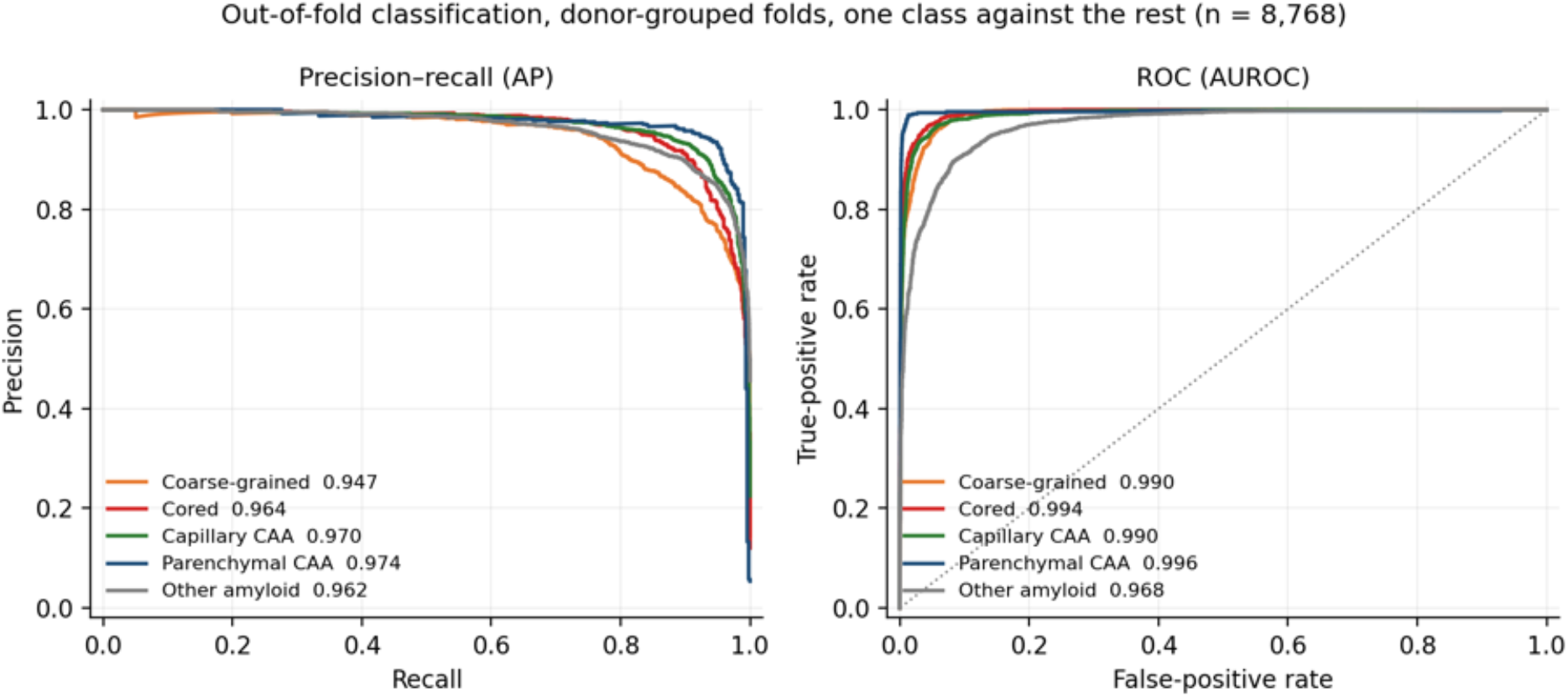
Out-of-fold classification with donor-disjoint folds, one class against the rest, on the averaged two-arm probability (8768 crops). **Left**, precision–recall, with average precision in the legend; **right**, receiver operating characteristic, with the area in the legend. Average precision ranges from 0.947 for coarse-grained plaques to 0.974 for parenchymal CAA.

**Fig. 4.**
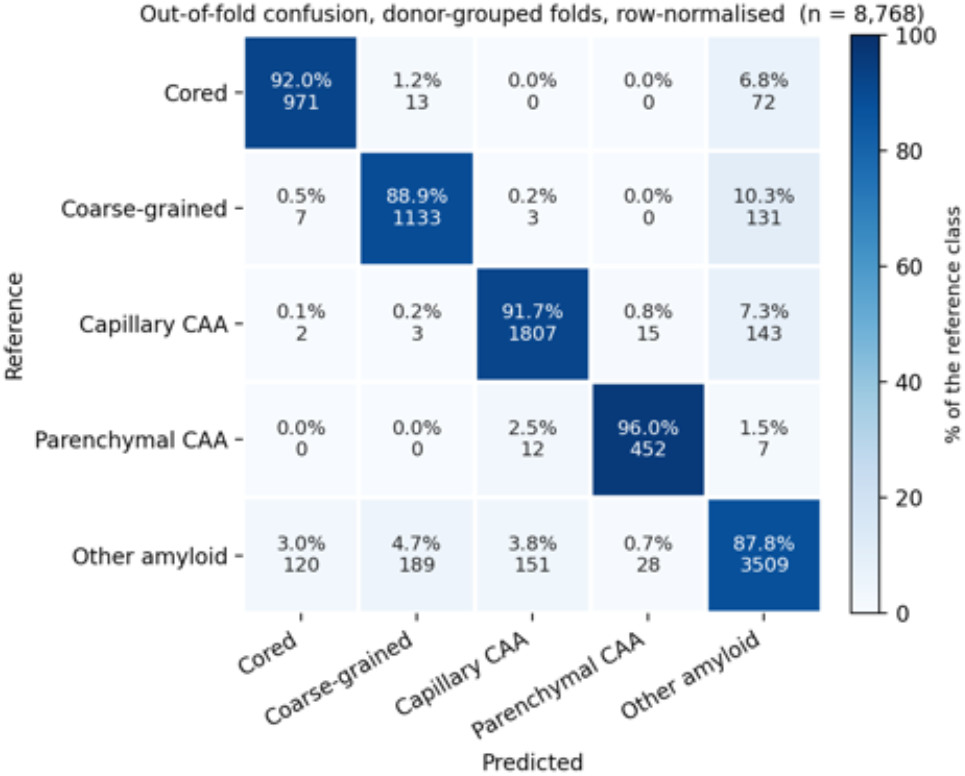
Out-of-fold confusion with donor-disjoint folds, row-normalized (each cell gives the percentage of the reference class, then the count; 8768 crops). Of 896 misclassifications, 93.9% involve the Diffuse/other category on one side or the other; confusion *among* the four amyloid classes accounts for 55 crops, 0.63 % of the dataset.

#### 3.2.3. The residual error is a taxonomic-boundary decision, not a subtype confusion

The five-class figure understates how well the four named morphologies are separated from one another. Decomposing the out-of-fold errors, 93.9% involve the Diffuse/other Amyloid boundary — between a named subtype and the residual Diffuse/other Amyloid category — a deposit assigned to a named type when it belongs to Diffuse/other Amyloid, or the reverse — and only 6.1% are confusions among the four amyloid classes (Fig. 4).

Stated as two nested problems: separating the four named morphologies from the residual Diffuse/other Amyloid category gives F1 0.913, whereas among crops whose reference and predicted labels are both one of the four named classes (n = 4.418), four-way macro-F1 is 0.985 (accuracy 0.988). The decomposition locates the weakest class more precisely. Among 1274 reference CGP crops, 1133 were classified correctly, 131 as Diffuse/other and 10 as another named class; among 1338 CGP predictions, 189 had the reference label Diffuse/other and 16 another named class. The CGP’s F1 of 0.868 does not reflect confusion with the other classes such as CCP/CAA1/CAA2, but rather the difficulty of deciding whether a given deposit meets the definition of a coarse-grained plaque, as opposed to being amyloid that falls outside the classes for which we are training. These are the same cases pathologists find challenging, highlighting the heterogeneity of amyloid pathology even within well-defined classes.

### 3.3. Comparison against published pipelines

We compared ViTamyloid against two published methods, Amyloid-YOLO (Wong et al. 2023) and PlaqueBox (Tang et al., 2019), on four reference standards drawn from four sources (Table 7). Every method was run at its published default configuration and scored by one matching rule, and detections compete only for reference objects of their own class, so a method is neither credited nor penalized for a taxonomy it does not train for. Each cell gives the area under the precision–recall curve and the best F1 along that curve, so no method is judged at an operating point its authors did not choose.

#### ViTamyloid leads on both classes on every standard

For cored plaques, the best F1 is 0.780 against 0.324 to 0.593 on the MCBB standard, 0.898 against 0.382 to 0.677 on the NBB standard, 0.746 against 0.570 to 0.618 on the UC Davis, UCLA and UC Irvine material, and 0.798 against 0.398 to 0.773 on the UC Davis ADRC hold-out. For CAA, it leads on all four (0.685, 0.773, 0.600, 0.727), although on the UC Davis ADRC hold-out the lead over PlaqueBox (0.571) rests on six CAA-positive crops, too few to separate methods (AUPRC difference +0.35, 95% CI − 0.50 to +0.54; best-F1 difference +0.16, − 0.33 to +0.47), in a set its original authors excluded from their CAA benchmark. The lead is significant (*p<* 0.05) in 26 of the 32 paired comparisons (Supplementary Table 11); it is not significant against PlaqueBox on the UC Davis ADRC hold-out (cored best-F1 difference +0.024, 95% CI − 0.109 to +0.158; CAA as above) or for CAA AUPRC on the UC Davis, UCLA and UC Irvine material against either comparator (+0.063, − 0.029 to +0.192 against Amyloid-YOLO; +0.048, − 0.072 to +0.207 against PlaqueBox). On the UC Davis, UCLA and UC Irvine set, scored on all 200 expert-reviewed tiles, 76 of ViTamyloid’s 161 CAA false positives at its best-F1 threshold, and 56 of Amyloid-YOLO’s 172, come from a single UC Irvine 6E10 slide whose 20 tiles show vessel-dense fibrous tissue with diffusely stained vessel walls. The experts disagreed on this slide: one of the four marked 93 CAA objects on it, against 17 in the consensus, and 56 of our 76 false positives fall on that expert’s marks.

#### The margin depends on the material being scored

On the two standards we annotated (NBB and MCBB), the nearest competitor reaches about three quarters of our cored F1 (0.677 against 0.898 on the NBB standard; 0.593 against 0.780 on the MCBB standard). Amyloid-YOLO trails by a similar margin on its own tiles (0.570 against 0.746), where PlaqueBox is closer in performance (0.618), but PlaqueBox nearly matches us on its own hold-out (0.773 against 0.798, not significant). The table therefore does not show a constant margin but instead shows that ViTamyloid is the only method that holds up across all four standards, which differ in antibody, scanner and image quality.

#### No comparator is consistently second

By best F1, the runner-up on cored plaques is Amyloid-YOLO on the MCBB and NBB standards and PlaqueBox on the UC Davis, UCLA and UC Irvine dataset (0.618 against 0.570) and on the UC Davis ADRC hold-out; the same split holds for CAA.

## 4. Discussion

This study introduces ViTamyloid, a computational pathology framework that converts amyloid *β* immunohistochemistry into continuous, lesion-resolved, and spatially explicit measurements. The pipeline distinguishes classic cored plaques, coarse-grained plaques, capillary cerebral amyloid angiopathy, and non-capillary parenchymal or leptomeningeal cerebral amyloid angiopathy, while retaining other A*β* positive deposits as a measurable category. Across complementary evaluations, ViTamyloid showed high detection precision, strong slide-disjoint and donor-disjoint classification, and better performance than published pipelines. The advance is therefore not simply automated detection, but scalable measurement of the composition and regional organization of A*β* pathology. These findings address the mismatch between the biological complexity of A*β* deposition and the intentionally simplified scores used in diagnostic practice. CERAD, Thal phase, and NIA AA criteria summarize plaque frequency and regional distribution, while consensus CAA protocols use ordinal regional scores (3–7). These systems remain essential for diagnosis and staging, but substantial variation in lesion burden and composition can exist within each category. Multicenter studies have also shown that plaque assessment varies with staining sensitivity, laboratory methods, and interpretation (8, 10, 11). ViTamyloid complements these standards by retaining continuous lesion number, area, density, class, and location. The technical results indicate that ViTamyloid gains most of its advantage through discrimination. Detection precision was 0.994 at the deployed operating point, and recall was 0.979 for deposits of 15 µm or larger. Donor-disjoint cross-validation produced a macro-F1 of 0.903 across five categories. Most residual errors occurred at the boundary between a defined lesion class and Diffuse/other Amyloid, with few mismatches among the four defined lesion classes. Most errors occurred at the boundary between a defined lesion and “Diffuse/other Amyloid”, with few mismatches among the four defined classes. ViTamyloid achieved the highest cored-plaque and CAA F1 on all four reference standards; Nominally significant differences at the unadjusted *p<* 0.05 level were observed in 26 of 32 paired comparisons. Earlier systems established that convolutional networks and object detectors can recognize cored plaques and CAA (38, 39), and large autopsy studies demonstrated the value of automated A*β* area measurements (40). ViTamyloid extends this work through pixel level segmentation, instance separation, artifact rejection, and a more informative lesion taxonomy. This work also illustrates the opportunity and practical difficulty created by the rapid evolution of foundation models. Keeping pace requires combined expertise in model selection, adaptation, dense prediction, calibration, validation, and neuro-pathologic phenotype definition. Foundation models are powerful starting points rather than complete solutions. In our ablation analysis, the adapted two-stream model outperformed frozen UNI2-h features (macro-F1 0.903 against 0.795), with the largest gain for classic cored plaques. This supports task specific adaptation that preserves broad representations while adding the spatial and morphologic information required for neuro-pathology. Software generalization across samples plays an important role in archival brain tissue. Histologic images vary across centers because of tissue processing, antibody selection, chromogen intensity, section quality, scanner optics, and resolution. Such domain shifts can reduce performance outside the development laboratory, and stain normalization alone does not consistently solve the problem (41–44). ViTamyloid was evaluated across independent collections generated with different A*β* antibodies and scanners. The small difference between crop-level folds, in which crops of one slide could fall on both sides, and slide- or donor-disjoint folds (macro-F1 0.910 against 0.898 and 0.903) suggests that classification relied primarily on lesion morphology rather than donor source. Because some external datasets combined all CAA into one class and lacked coarse grained plaque labels, external validation of every subtype remains an important next step.

## 5. Conclusion

The largest scientific opportunity is the application of lesion-resolved quantification to brain banks and longitudinal clinico-pathologic cohorts. These resources link donated tissue to years of cognitive testing, clinical phenotyping, genetics, imaging, and fluid biomarkers (36, 37). Autopsy studies already show that neuro-pathologic burdens and subtypes relate differently to cognitive trajectories (58–60). Continuous regional measurements could reveal why individuals assigned the same Thal phase or CERAD category follow different clinical courses and identify lesion combinations associated with resilience, rapid progression, or specific phenotypes. Autopsy is cross sectional and cannot directly establish temporal sequence within one individual. Integration with longitudinal antemortem data can nevertheless support population level models of disease trajectories. Processing 1,630 whole-slide images demonstrates the feasibility of applying ViTamyloid at this scale. The framework can also sharpen correlations with imaging and fluid biomarkers. Within sampled tissue, autopsy histology is the microscopic ground truth for lesion identity, vascular compartment, cortical layer, and location. Amyloid PET and fluid biomarkers perform well for identifying substantial AD neuropathologic change, but discordance occurs, and PET sensitivity depends on histologic density and distribution (21–25). Lesion resolved histology could determine which plaque and vascular patterns contribute to PET signal, explain biomarker discordance, and characterize sparse residual pathology that may not produce a positive scan.

This capability is particularly timely in the treatment era. Lecanemab and donanemab trials demonstrated substantial lowering of amyloid PET signal and modest slowing of clinical decline, while identifying amyloid related imaging abnormalities as an important safety outcome (18, 19). As more treated individuals come to autopsy, neuropathology will be essential for determining which lesion classes were cleared, which persisted, and whether A*β* was redistributed among cortical layers, plaques, capillaries, and larger vessels. AN1792 autopsies showed extensive but regionally variable plaque removal, persistence of downstream pathology, and continued or increased vascular amyloid (29–31, 33, 34). A recent aducanumab autopsy likewise demonstrated regional, laminar, gyral, and sulcal heterogeneity in residual A*β* (28). ViTamyloid could potentially convert these observations into comparable quantitative endpoints linked to drug exposure, ARIA, imaging, fluid biomarkers, and clinical response. Several considerations define the next validation steps. Reference labels reflect expert interpretation of a morphology based taxonomy, and the weakest class, coarse grained plaque, was limited mainly by its boundary with Other Amyloid. Multireader adjudication and direct measurement of interobserver agreement will clarify the attainable ceiling for each class (7, 8). External studies should include prospective annotations for all classes, broader disease stages, varied tissue quality, additional antibodies and scanners, and treated brains. Thresholds should also be selected separately for lesion counting and total A*β* area. Automated Thal and CERAD summaries require fully sampled brains and direct validation against recorded scores. A*β* immunohistochemistry alone cannot establish the neuritic component required for CERAD plaque scoring (3, 6). These steps will define the appropriate roles of ViTamyloid in discovery studies, clinical trials, and standardized diagnostic assistance. Overall, ViTamyloid advances automated A*β* analysis from broad deposit detection toward portable, lesion resolved pathomics. Its combination of technical accuracy, biological specificity, and whole slide scalability creates a route for connecting microscopic A*β* heterogeneity with genetics, clinical trajectories, biomarkers, imaging, and therapeutic response.

## Data Availability

All data produced in the present study are available upon reasonable request to the authors

## 6. Data Availability

Partial data collections utilized in this study are already publicly accessible. The annotated image dataset from Wong et al. is freely available via the Open Science Framework at `DOI: 10.17605/OSF.IO/FCPMW`. The raw whole-slide images and annotated plaque-level datasets from Tang et al. are openly hosted on Zenodo at `DOI: 10.5281/zenodo.1470797`. External validation data sourced from the Netherlands Brain Bank (NBB) are accessible according to their respective institutional repository guidelines. Access to the Mayo Clinic Brain Bank (MCBB) dataset is restricted; these data will be made available upon reasonable request, subject to data use agreements and official clearance from the Mayo Clinic.

## ACKNOWLEDGEMENTS

This work was supported by the Mayo Clinic in Florida, Jacksonville, FL, through the Brain Bank Modernization funding project by the Department of Neuroscience and NIH grants K24AG053435, R01AG070826, R01AG075802 and P30AG062677. Ethical oversight was provided by the Mayo Clinic Institutional Review Board (IRB #15-009452). This study used deidentified archival postmortem tissue, histologic images, and associated data. The Mayo Clinic determined that this activity did not constitute human subjects research [insert exact institutional determination and ID].

## Appendix I

Figure 5 illustrates representative image crops extracted from the dataset compiled by Tang et al. (2019), showcasing various instances of potential plaque candidates encountered during neuropathological evaluation. These visual samples highlight the morphological variability and structural features of candidate plaques within tissue scans, offering a qualitative view of the raw visual patterns used for subsequent detection and classification tasks.

**Fig. 5.**
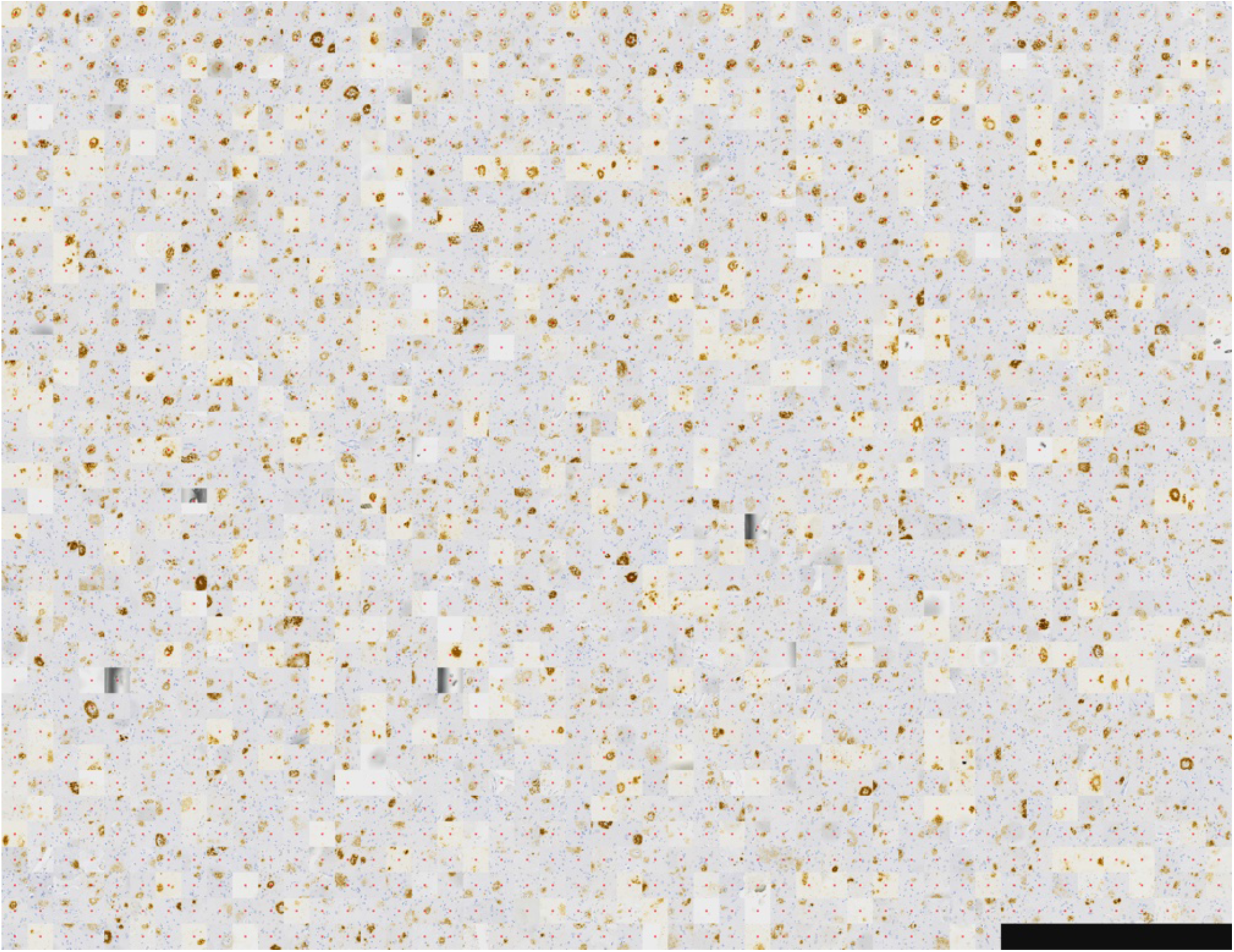
Example of image crops extracted from data collection in Tang et al. (38), containing plaque candidates.

Table 9 expands on the classification performance summarized in Table 6 across 8,768 crops (with classes assigned by the argmax of the averaged probability), providing 95% confidence intervals for the primary donor-disjoint cross-validation alongside two five-fold sensitivity analyses: slide-disjoint folds and antibody-stratified crop-level folds. All intervals and paired differences are derived from 2,000 bootstrap replicates that resample donors—keeping all slides of a given donor from MCBB dataset#1 (6E10 or 6F3D) together—to ensure a direct and robust comparison across all three validation designs.

Table 10 presents the comprehensive, granular performance breakdown that complements the summary provided in Table 7. To account for hierarchical data dependencies, the reported 95% confidence intervals are derived from 2,000 bootstrap replicates that resample at the level of the independent unit (donor or slide) while evaluating every method on identical matched detections. Precision and recall are assessed across distinct confidence thresholds, offering full transparency into model behavior across the reference classes.

Table 11 details the quantitative performance differences between ViTamyloid and each comparator across all metrics in Table 7. Derived from the same 2,000 paired bootstrap replicates as Supplementary Table 10, these comparisons include two-sided *p*-values calculated as twice the proportion of replicates on the opposite side of zero (unadjusted for multiple comparisons, with a minimum reporting threshold of*<*0.001).

